# ProBioQuest: a database and semantic analysis engine for literature, clinical trials, and patents related to probiotics

**DOI:** 10.1101/2022.02.17.22271109

**Authors:** Po Lam Chan, Susana Lauw, Ka Lee Ma, Nelson Kei, Ka Leong Ma, Yiu On Wong, Ho Yan Lam, Rina Ting, Tsz Kwan Yau, Wenyan Nong, Dandan Huang, Yichun Xie, Peter Chi Keung Cheung, Hoi Shan Kwan

## Abstract

The use of probiotics to improve health via the modulation of gut microbiota has gained wide attention. The growing volume of investigations of probiotic microorganisms and commercialized probiotic products has created the need for a database to organize the health-promoting functions driven by probiotics reported in academic articles, clinical trials, and patents. Currently, no such database is available. We constructed ProBioQuest to collect up-to-date literature related to probiotics from PubMed.gov, ClinicalTrials.gov, and Patents View. More than 2.8 million articles were collected by the end of 2021: PubMed.gov: 2 656 818; Clinical Trials.gov: 205 349; Patents View: 32 536. Automated information technology-assisted procedures enabled us to collect the data continuously, providing the most up-to-date information. Statistical functions and semantic analyses are provided on the website as an advanced search engine, which contributes to the semantic tool of this database for information search and analyses. The semantic analytical output provides categorised search results and functions to enhance further analysis. A keyword bank is included which can display multiple tables of contents. Users can select keywords from different displayed categories to achieve easy filtered searches. Additional information on the searched items can be browsed via the link-out function. ProBioQuest is not only useful to scientists and health professionals, but also to dietary supplement manufacturers and the general public. In this paper, the method we used to build this database-web system is described. Applications of ProBioQuest for several literature-based analyses of probiotics are included as examples of the various uses to which this search engine can be put. ProBioQuest can be accessed free of charge at http://kwanlab.bio.cuhk.edu.hk/PBQ/.

**Database URL:** http://kwanlab.bio.cuhk.edu.hk/PBQ/

## Introduction

The use of probiotics to improve health via the modulation of gut microbiota has gained wide attentions (1). Probiotics have been defined as “live microorganisms which when administered in adequate amounts confer a health benefit on the host” (2). Various health conditions such as obesity, type 2 diabetes, cancers, gastrointestinal disorders, and depression might be alleviated through probiotic supplementation (3). At present, *Lactobacillus* and *Bifidobacterium* are the major lactic acid bacterial genera used in the probiotic supplements. The genus *Lactobacillus* has been reclassified into 25 genera, which include the *L. delbrueckii* group, *Paralactobacillus*, and 23 novel genera (4). The growing volume of investigations of probiotic microorganisms and commercialized probiotic products has created the need for a database to compile the health-promoting functions driven by probiotics reported in academic articles and clinical trials.

Online databases can be created to gather data and provide hyperlinks, enabling users to access information of interest quickly and effectively (5). To date, several imperfect databases with the theme of probiotics have been developed. Previously, information on the functions of probiotics that were marketed or used in clinical, field, and animal studies were provided in a probiotics database named PROBIO (6). Additionally, the functions of probiotics originating from fermented foods could be accessed using the probiotics database PBDB (7). Unfortunately, PROBIO and PBDB are no longer accessible. The probiotics databases AEProbio (http://usprobioticguide.com/) and Optibac Probiotics (http://www.optibacprobiotics.com/uk/professionals/probiotics-database) are currently accessible and can be used free-of-charge. Although their target users are health professionals, these databases are not updated frequently, and the scope for searching remains limited. PROBIO and PBDB contained 997 and 1730 probiotics, respectively (6,7). However, only 21 probiotic strains were included in the Optibac Probiotics, a remarkably low figure compared to the coverage of PROBIO and PBDB. The primary search entry of Optibac are probiotic strains while that of AEProbio was health conditions. Users were limited to performing unidirectional searches using these two databases. No tools are provided by either database’s web interface to facilitate further searching from the output. In addition, it was not possible to search for safety information regarding probiotics in humans in these databases.

As the growing need for detailed information is not presently being satisfied, we have established a new database, ProBioQuest, to collect up-to-date articles related to probiotics from PubMed.gov, ClinicalTrials.gov, and PatentsView. Automated information technology-assisted procedures have enabled us to collect the data continuously. Statistical functions are provided on the website, an advanced search engine, which contributes to the semantic tool of this database for information search and analyses. The output provides categorized search results and functions for further searching. ProBioQuest includes a keyword bank, which displays multiple tables of contents containing concepts of probiotics, microbiome, diseases, and food. Users can select keywords from different displayed categories, enabling them to conduct a customized search without difficulty. Additional information on the searched items can be browsed on Google Image, Google Scholar, Wikipedia, Drugs.com, Amazon, Drug Bank, Google Trend, and Disbiome via the link-out function, giving ProBioQuest a clear superiority over existing knowledge-based search engines for probiotic studies. Therefore, ProBioQuest can not only be utilized by scientists and health professionals, but also dietary supplement manufacturers and the general public. In this paper, the method we used to build this database-web system is described. We also list source websites and provide instructions for using the database.

## Methodology

ProBioQuest is a database-web system with an advanced search engine designed for online intelligent searching of probiotics-related articles. Figure 1 shows the workflow of ProBioQuest. Targeted information from 3 public web resources was collected to create this database. With the aid of Application Programming Interface (API) technology (26), the database was developed by automated data collection using Perl script, which enabled later data processing. The comprehensive keyword lists allowed searching of in-depth probiotics-related resources. In the probiotics-related keywords bank generation process, the number of occurrences of each keyword was counted, with the incorporation of each article into the MySQL (27) database with statistical analysis. After data collection, Solr (28) was used for indexing and building a searchable probiotics database website, including back-end big data analytic applications. Keyword frequency was counted and displayed for easy visualization of relative popularity. A visual analytic, the “Keyword Cloud”, appeared on the front page. This provided a summary of the database by using the number of occurrences for each keyword to compute its corresponding size. For the user interface, we used JavaScript as the scripting language to build the website, exploiting its statistical functions for search results.

**Figure 1.**
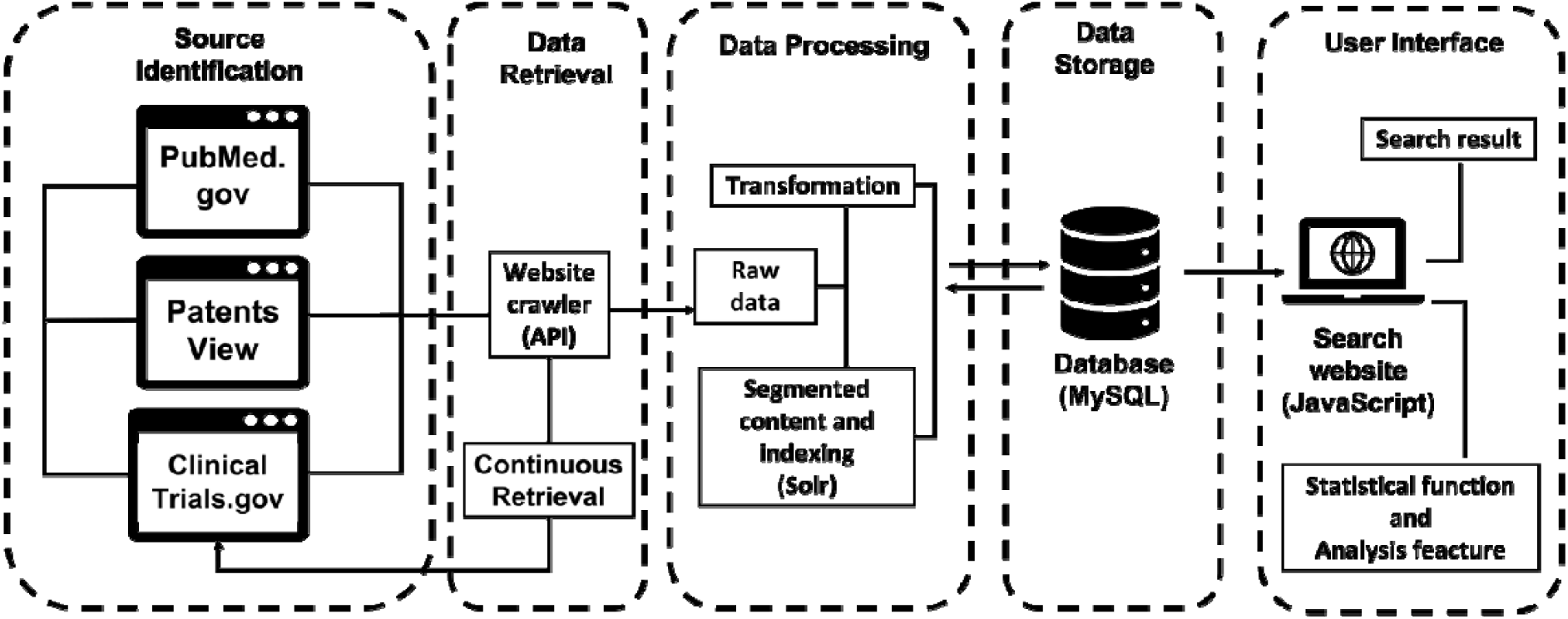
The workflow of ProbioQuest. The database works with continuous data retrieval from 3 public sources via API function. Our in-house Perl script transforms the raw data into the desired structure and then inputs it to MySQL. Subsequently, Solr creates segments of the content and indexes according to the title and abstract or summary of the processed data. The user interface was built by using JavaScript for searching, with its statistical function and analysis feature.

### Source identification

ProBioQuest collects relevant information from various sources, including journal articles, patents, and reports of clinical trials. The source websites we used were PubMed.gov (https://pubmed.ncbi.nlm.nih.gov/), PatentsView (https://www.patentsview.org/), and (ClinicalTrials.gov (https://clinicaltrials.gov/). Our database extracted targeted information from these websites, which provide a wide coverage of the corresponding type of resource. These three websites are generally considered to be reliable and professionally managed, and were therefore treated as sources of trustworthy information in our development of ProBioQuest.

The first website, PubMed.gov, contains more than 32 million abstracts and citations related to life sciences, biomedical science, chemical sciences, behavioral sciences, and bioengineering. PubMed.gov includes articles that are in progress, not yet officially published, collected or not selected by the Medline database, published by National Institutes of Health (NIH) subsidized researchers, and every document held by the National Library of Medicine (NLM) and the National Center for Biotechnology Information (NCBI). With the inclusion of the comprehensive academic literature database provided by PubMed.gov as a source, ProBioQuest can extract an extensive amount of information for exploitation by professional users in searching for academic articles.

The second website, ClinicalTrials.gov, is a U.S. government website maintained by the NLM at the NIH. It is updated daily and provides the most recent information about clinical research studies. This website contains information of over 403 000 clinical studies and research studies conducted around the world. It consists of a Protocol Registration and Results System (PRS), which is a web-based data entry system. Its public clinical results databases provide a route for extracting biomedical outcomes. Integrating ClinicalTrials.gov into our database enables health care professionals to discover the latest probiotics-related investigations on the ProBioQuest website.

The third website, PatentsView, allows users to search patenting activities around the U.S. Exploring data among 5 million U.S. patents, 1.2 million inventors, and 250 000 assignees data in 40 years, and extracting the disclosed patents in PatentsView allows ProBioQuest to include up-to-date probiotics-related patents. This makes for an efficient and easy patent search, enabling probiotics development experts to monitor market trends.

### Data extraction, processing, and storage

This database used the technique of API to extract all information from the source website databases for information completeness. We executed the scraping among sources of websites. Data records were searched and filtered out by keywords extracted. We used “Bacteria”, “Probiotic”, “Gut microbiome”, “Gut microbiota”, “Disease with gut”, and “Health with gut” as search terms to retrieve the data. As we used Perl language to automate manual workflows, the computer performed the monotonous work of screening, filling, and uploading. Rules and fields were set, then targeted information was extracted, including title, journal type (if from PubMed.gov), source, abstract or summary, authors or inventors or researchers, publication date, and URL. Each extracted record was converted into columns and rows in the database by Perl script. Data flowed from the source websites to our ProBioQuest MySQL database. The collected data were stored in MySQL as a temporary database, integrating all the information required for the ProBioQuest database website.

After data extraction, the open-source software Solr was used to index the entries in the MySQL database. Abstract search and near real-time indexing were established by Solr. Indexing, a process of turning the original data into a highly efficient cross-reference lookup to facilitate rapid searching, is then performed. Solr does not index text directly, but rather breaks the text into individual tokens, then consults the search index. Matching results are retrieved, then sorted in the requested order (e.g. publication date). The purpose of storing an index is to optimize speed and performance in finding relevant documents for a search query. With an index, a search engine does not need to scan every document in the database, economizing on machine time and computing power.

Data extraction was maintained according to the keyword bank, and included technical phrases of bacteria, probiotics, gut microbiome, gut related disease and health. The design of the keyword bank was based on our previous database “Food Safety Information Database for Greater China” (23). We collected terms and bacteria names from “NCBI Taxonomy” (24) and “Clinical Guide to Probiotic Products Available in USA” (25). “NCBI Taxonomy” is a comprehensive update resource with a list of accurate organism names and classifications (24). Thus, we used “NCBI Taxonomy” to find probiotic bacteria and microbiome related to gut, and further screened the bacteria name. The “Clinical Guide to Probiotic Products Available in USA” provides a list of probiotics. We construed and separated the words and terms in this list into more specific categories. To provide the correlation with searched keywords, the frequency of these keywords was calculated.

### Search procedure and advanced search function

The scalable and interactive functions of ProBioQuest were developed by JavaScript. One of the products was ProBioQuest’s keyword cloud (Figure 3A). Inside a keyword cloud, each keyword size is directly proportional to its frequency of occurrence. Showing keywords in different sizes can help users to choose which keywords to use. As JavaScript allows interaction between websites and users, clickable buttons can be made by coding. Users can tap the appropriate key on their keyboard to conduct searching instantly. ProBioQuest’s keyword bank is another useful function. Words in both the keyword bank page and sidebar (e.g. “add keyword”, “delete keyword”, “direct search”) enable searches to be refined. In “Bacteria/Probiotics” and “Microbiome” category, keywords can be searched externally in BacDive (29) and NCBI for genome and taxonomy. These functions allow users to customize their search results and provide more accurate search results suited to the needs of different users. Finally, users can use the output function of the searched results and keywords. There is an export function which allows users to extract what they request as a spreadsheet (.csv) for their reference.

Besides offering keyword-related functions, ProBioQuest also allows users to link with external websites. Since only the abstracts of journal articles or clinical trials have been captured, users can click on the titles, which are linked to their original websites. Users can obtain full-text information from selected articles by this function without being overwhelmed with excessive information or overloading the database website. To complete more advanced searching, there is an additional information section on the sidebar. ProBioQuest integrates popular and useful search engines such as Google Image, Google Scholar, Wikipedia, Drugs.com, Amazon, Drug Bank, Google trend and Disbiome. By clicking the corresponding button on the sidebar, users can search the keyword on any or all of their websites, for a customized searching journey.

## Results and discussion

### Description of ProBioQuest, key features and applications

ProBioQuest was constructed as a database-semantic analysis engine platform with the link http://kwanlab.bio.cuhk.edu.hk/PBQ/. It is intended for bioinformatics of probiotics for researchers, healthcare professionals, probiotics developers and the general public to query and analyze probiotics information in relation to microbiome, health conditions, and foods. This platform offers a scientific overview in the form of tables of contents of search terms and insights of relevant key concepts. Users can also explore by selecting from a keyword cloud displayed on the homepage. Further information about bacteria features, marketed probiotics, drugs, and its public awareness over time are available by external links. A total of 2 894 703 articles were collected by the end of 2021: PubMed.gov: 2 656 818; Clinical Trials.gov: 205 349; PatentsView: 32 536 (Table 1.). ProBioQuest is set for continue retrievals and analyses of the literature automatically at fixed time intervals.

**Table 1.** Breakdown by source of entries in the ProbioQuest database (Data accessed on 31st December, 2021).

|  | **Number** |
| --- | --- |
| Total entries in the database | 2 894 703 |
| Entries from Pubmed.gov | 2 656 818 |
| Entries from PatentsView | 32 536 |
| Entries from ClinicalTrials.gov | 205 349 |

On the homepage, users can select from a keyword cloud constructed from the most frequent keywords from articles. Alternatively, they may input the relevant keyword in the search bar. Suggestions based on our keyword bank will pop up while the user types. Each query, by default, searches from all sources in our database according to the title and abstract of sources and returns previews of articles, with the most relevant listed at the top in “Search Result”. In the preview, users can find title, author names, relevant sentence(s), source, and publication date. For clinical trials, users can click “Show Details” for identifier, study type and location. Users who wish to read further are directed to an article by a simple click on its title.

In “Keyword Bank”, users can find the novel feature of ProbioQuset for keyword or key concept discovery on interested topics. This tab shows the statistical analysis of relevant keywords and lists in descending order of number of articles they co-existed. Keywords are divided into categories. These keywords can also be found in the scrollbar on the left of the screen. Secondary keywords can either be appended to the search, or excluded from it, or individually searched, by simply clicking “Append”, “Exclude” and “Search” respectively. Additionally, the keywords of “Bacteria/ Probiotics” and “Microbiome” can be linked to the NCBI Genome List, the NCBI taxonomy database or the database BacDive for further information of these entities by selecting “Genome”, “Taxonomy”, or “BacDive” respectively. Additional external links (e.g. Google image) displayed at the end of the left scrollbar are directed to search results of the same keyword(s) used in ProBioQuest for easy access. Search results in ProBioQuest can be downloaded in the “Export” tab. Figure 2 shows window captured examples of semantic analysis and exploration on the user interface of ProBioQuest. Moreover, user guides are provided on the interface and a YouTube video has been created to demonstrate the search key points in ProBioQuest.

**Figure 2.**
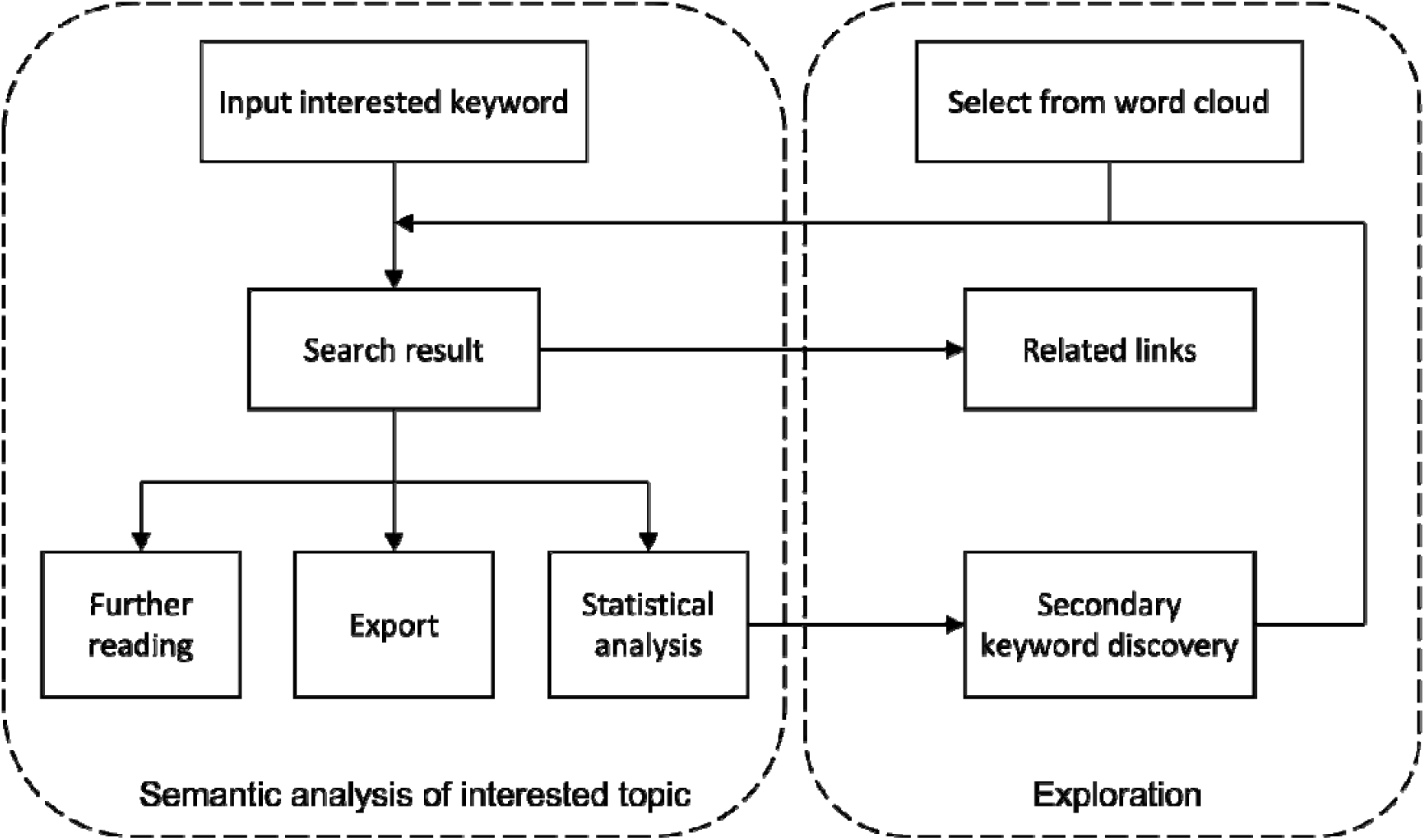
Flow chart of semantic analysis and exploration on the user interface of ProBioQuest. Users can simply input the keyword of interest, and ProBioQuest will show related search results. Users can then click the results for further reading or export the selected result. To explore deep searching, ProBioQuest provides a statistical analysis of the search result and Keywords Bank allows further searching by adding a secondary keyword.

ProBioQuest is not only an organized database but also a semantic tool to help users to search by disease, disorder, and symptoms for overall concept of the health or diseases condition, and potential probiotics to use. Below is a demonstration using “autism” as an example. Example questions: “Are there any conditions commonly experienced by autism patients?” and “Have any probiotics been tried to help autism?” Users could search the word “autism” and the result returns 2 294 articles (Figure. 3A&B). Clicking the search result articles will lead to the sources. A clearer understanding of autism can therefore be achieved by reading the articles. “Keyword Bank” tab provides the overall concept that autism is commonly discussed with other conditions such as anxiety, schizophrenia, hyperactivity and dysbiosis in the “All Keywords” category (Figure. 3C). For users interested in finding probiotics beneficial to autism patients, the keyword category “Bacteria / Probiotics” offers the relevant information quickly. The potential probiotics targets are rapidly narrowed to 10 species and 3 strains. The top species found is *Lactobacillus plantarum*. It can be linked out for its related taxonomy and strain information such as culture conditions (Figure. 3D). By appending *Lactobacillus plantarum* to the search, a species strain and clinical trial of this particular strain is found. Its potential in relieving autism can be further evaluated. For further information in respect of the searched keywords, our database-website system directly links out to search for marketed probiotics products on the left scroll bar, or exports the search result as a CSV file. (Figure. 3E). A more detailed discussion on the bioinformatic insights obtainable from searching “*Lactobacillus rhamnosus* GG” is provided below in 4.3.

**Figure 3.**
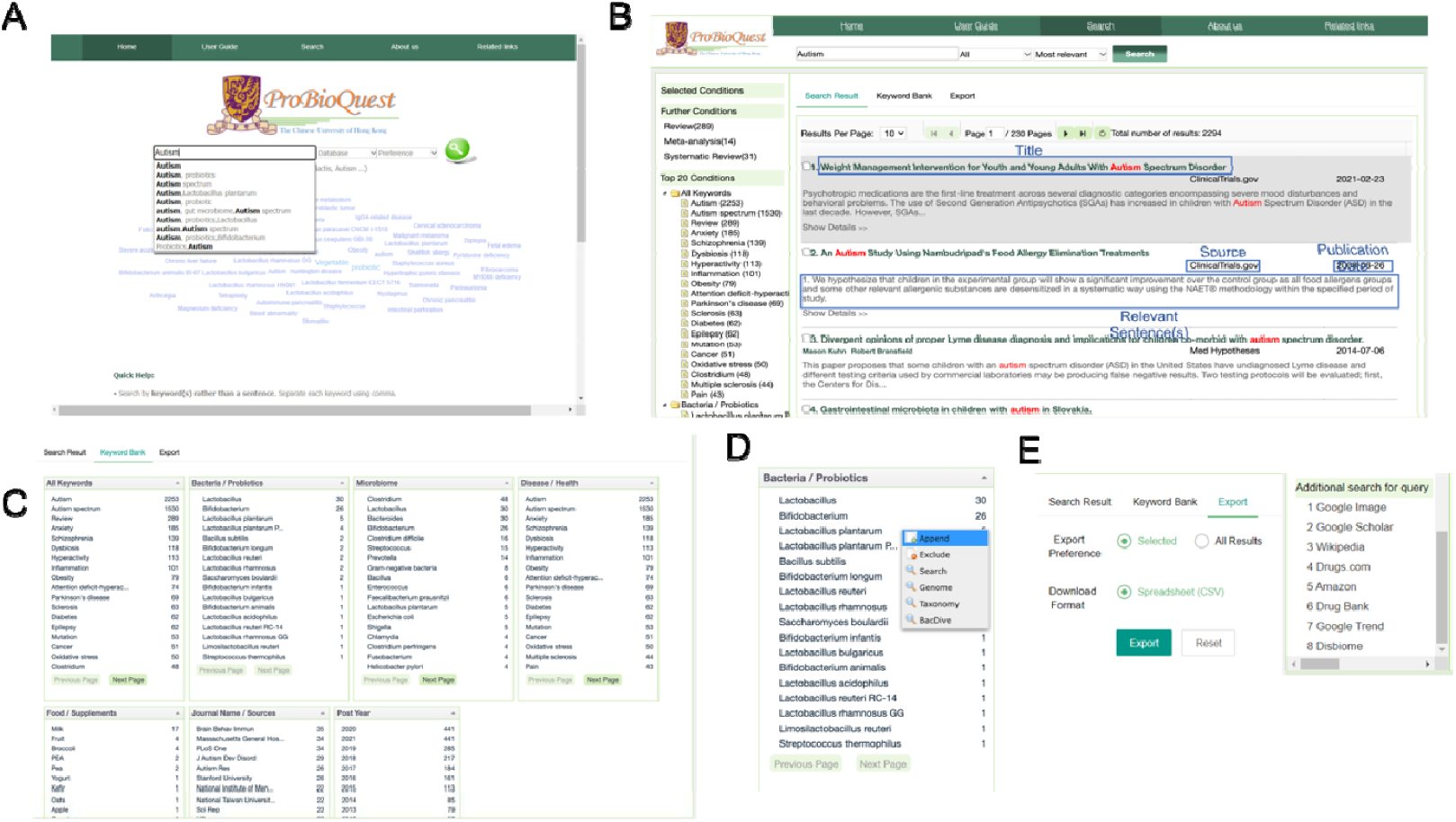
An Overview of the user interface of ProBioQuest. (A) Home page includes a keyword cloud of the most frequent keywords from all seed articles, a search bar which offers suggestions while the user is typing, and a help box for users unfamiliar with the system’s details. (B) Autism as an example of input and the search result. Each article is linked out to ClinicalTrials.gov, PubMed.gov or PatentsView with a simple click. (C) Novel feature in Keyword Bank tab. This tab shows relevant keywords in descending order of number of articles they co-existed, which aids secondary key concept discovery of the interested topic. All keywords are divided into categories. (D) The secondary keyword can be appended into the search bar or linked out. (E) Search result can be exported as CSV format or linked out to search for additional information.

### Evolving research interests of probiotics as revealed by ProBioQuest

We demonstrate here how the Keyword Bank feature in ProBioQuest can help users to gain insights into research trends of probiotics functions. We entered the word “probiotic; probiotics” into the search box, which displayed 28 561 articles in the “Search Results” tab. In the “Keyword Bank” tab, the number of articles in each year was shown in “Post Year” in descending order of hits. The number of articles on probiotics has been growing rapidly (Figure. 4). The year can be appended into our search. We appended the year 2021 and clicked “Keyword Bank”. We collected all items and the corresponding frequency present on the first page of the “Disease/Health” category under Keyword Bank. We conducted this data collection from 1991 to 2021. From the list of 498 items, 95 unique Disease/Health items were found. We obtained the top 30 items by ranking them in terms of their frequency from 1990 to 2021. The data were organized into groups of 5 years to show the changes in the numbers of returns (Figure. 5).

**Figure 4.**
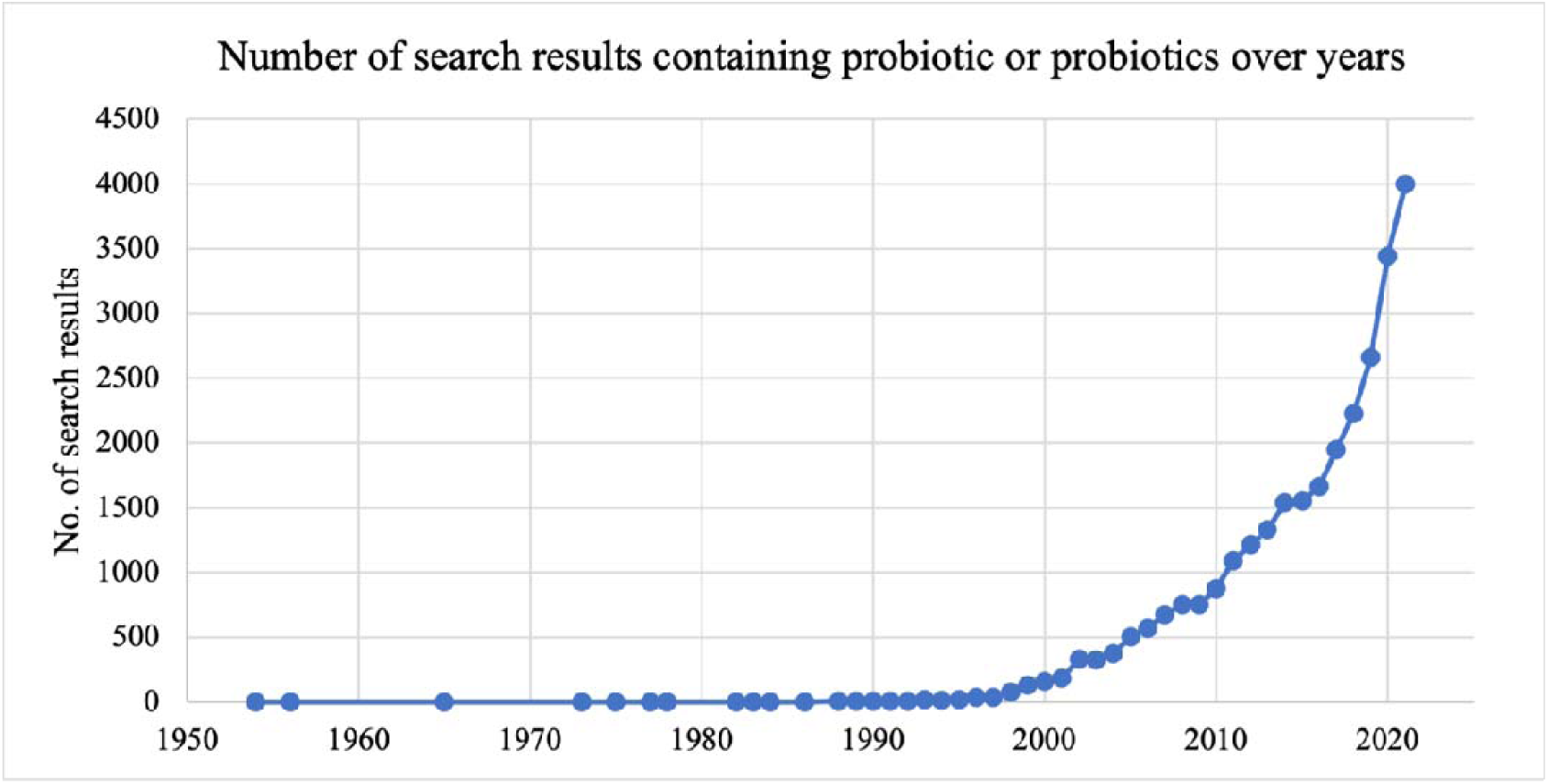
The number of articles containing “probiotic/probiotics” from 1950 to 2021. The number of articles on probiotics has been growing rapidly during the past two decades.

**Figure 5.**
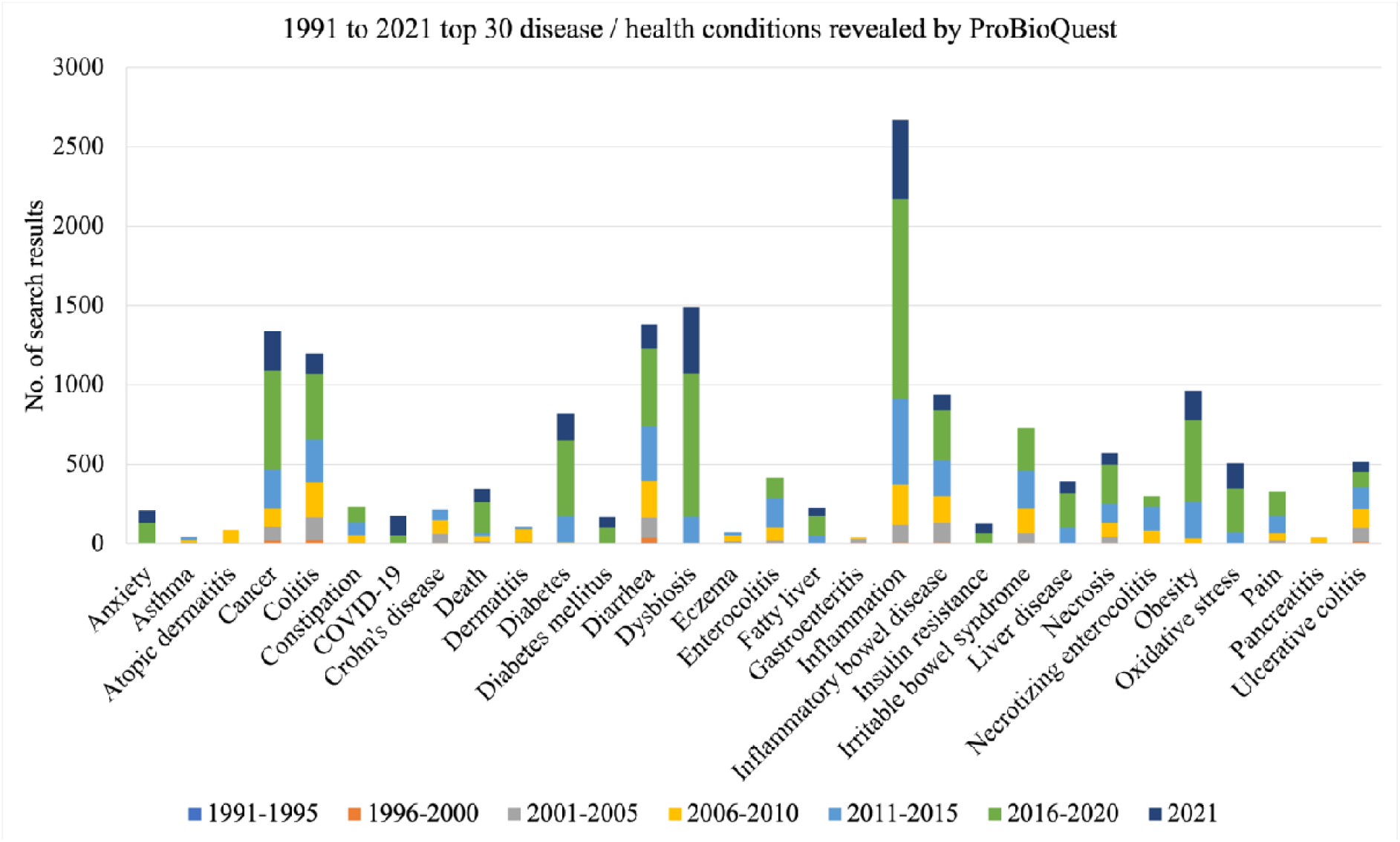
The number of articles of top 30 Disease/Health in 5 years groups. Popular topics such as inflammation, diarrhea and cancer maintained a high rate of occurrence in the articles from 1991 to 2021.

The top 10 most popular Disease/Health keywords in 2020 were selected for further analysis of their trends from 2001 to 2020 (Figure. 6). Inflammation and dysbiosis have clear trends of great growth in numbers. Specific diseases of awareness on metabolic disorders like obesity, diabetes, and oxidative stress also increased in the past ten years, though to a lesser extent. Researchers have shifted their interest from probiotics functions in gastrointestinal issues like colitis, inflammatory bowel disease, diarrhea and constipation to more physiological conditions. Almost all results on anxiety come from the past 5 years, indicating a rise in psychobiotics research. We also found that researchers sometimes use more than one term for the same health condition. The number of articles on atopic dermatitis and eczema, for example, were counted separately, even though they are two ways of describing the same skin condition. Unifying terms will benefit researchers in the future when reviewing published articles.

**Figure 6.**
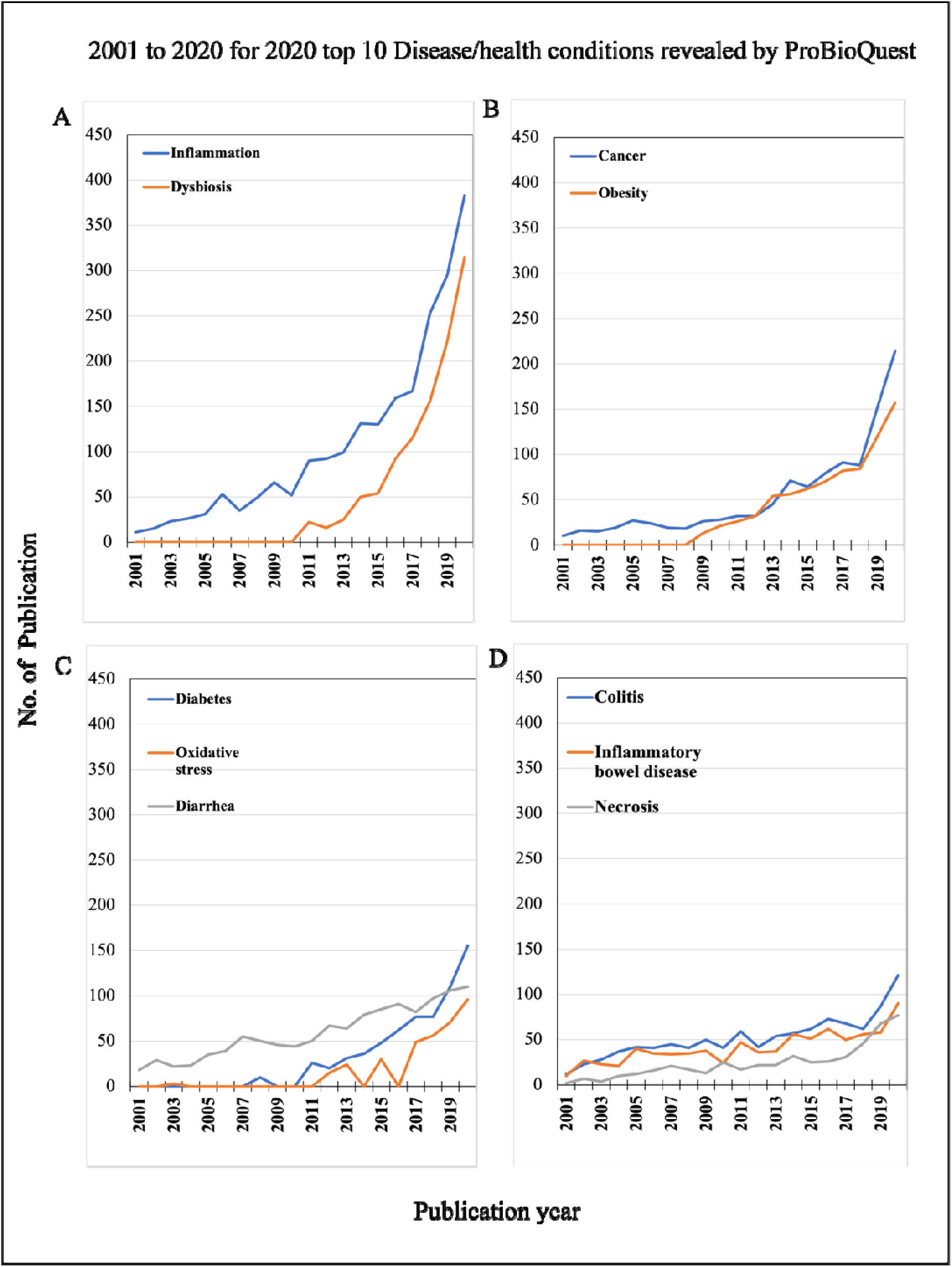
2001 to 2020 for 2020 top 10 Disease/Health conditions revealed by ProBioQuest. The top 10 most popular Disease/Health keywords in 2020 were selected for further analysis of their trends during the two decades from 2001 to 2020. (A) Inflammation and dysbiosis have clear trends of marked growth in numbers. (C-D) Specific diseases of awareness on metabolic disorders like obesity, diabetes and oxidative stress, and diseases of abnormal cell growth like cancer and necrosis also increased in the past ten years, although to a lesser extent.

### Use of ProBioQuest to gather comprehensive and up-to-date information of a probiotic strain: *Lactobacillus rhamnosus* GG as example

As an example of how comprehensive and up-to-date information on the probiotic strain *Lactobacillus rhamnosus* GG (LGG) can be gathered, we used ProBioQuest to answer the following three questions: (1) What is the most popular health condition associated with the probiotic strain *Lactobacillus rhamnosus* GG (LGG)? (2) What probiotic bacteria are associated with LGG? (3) What probiotic bacteria and foods are associated with LGG? The users could enter the word “*Lactobacillus rhamnosus* GG” into the search box, which will display a total of 929 items in the “Search Results” tab of the data page. Variation of the item number and frequency statistics would be observed at different enquiry times owing to the continuous updating of ProBioQuest. To narrow down this vast amount of information to the more relevant subsets, users could click items on the “Keyword Bank” section on the data page. It has the frequency ranking of all keywords, probiotics, microbiome, diseases, food, sources, and post years. Inflammation (n=99) is the most frequently mentioned health condition in the literature, and 2020 (n=101) was the year when most of the research articles related to LGG were published. Bifidobacterium is an important member of the microbiome, which ranks first both in the “Bacteria / Probiotics” category and in the “All Keywords” statistics. Adding “Bifidobacterium”, by choosing the “Append” option, will decrease the total number of results from 929 to 108 results in two clicks. To address the third question, users could return to the Keyword Bank to append “Milk” under “Food / Supplements”, respectively. The total number of results related to *Bifidobacterium*, milk and LGG were narrowed down to 15, demonstrating the ease with which customized search for the information of a probiotic strain can be conducted using ProBioQuest (Figure7).

**Figure 7.**
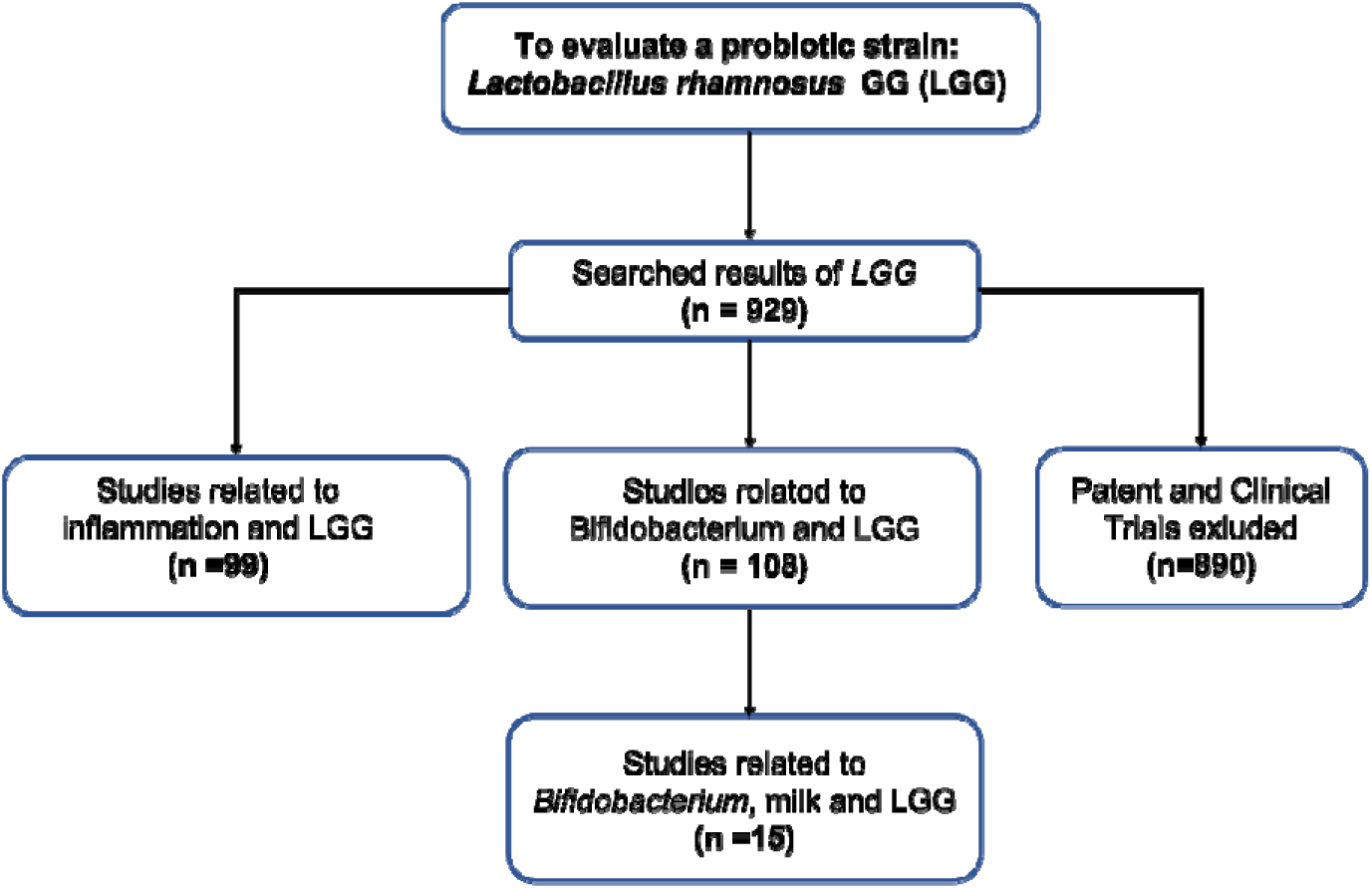
Flowchart of using ProBioQuest to evaluate a probiotic strain-*Lactobacillus rhamnosus* GG. Searching *Lactobacillus rhamnosus* GG. in ProbioQuest returned 929 results. There were 890 articles from Pudmed.gov. To narrow down the number of results to those of more specific interest, a second keyword (“*Bifidobacterium*” or “inflammation”) was added. This reduced the number of results to 108 and 99 respectively. The addition of more keywords would further reduce the number of results and narrow the field of enquiry.

### Emerging application of probiotics revealed by ProBioQuest: Autism spectrum disorder

High relevance of results is one of the key ways in which ProBioQuest can assist users in acquiring desired information efficiently. In recent years, probiotics have been proposed to play a role in treating autism spectrum disorder (ASD), not only to alleviate gastrointestinal dysfunction but also to reduce the severity of ASD. To investigate the possible role for probiotics, a review of the therapeutic effect of probiotics on ASD is required. ProBioQuest, with data collected from a variety of reliable sources, can provide such information. When we used the keywords “probiotic” or “probiotics” and “autism” with Database “All” and Preference “Most relevant” to search, the page of this entry displayed a total of 164 results, all of which were relevant to ASD and probiotics, from 2002 to 2021. That topic of studies gradually increased from 2018.

On the tab of “Keyword Bank”, ProBioQuest offers useful ideas relevant to the searched topic. Seven columns of keywords are offered for selection to further query targeted information about the associations between autism and probiotics, as well as sorted out diseases or specific probiotics that would help us to further study ASD and probiotics. In the Bacteria/ Probiotics column, there were 2 genera *Lactobacillus* and *Bifidobacterium* together with 5 species and 3 strains of probiotics used in studies that were mentioned in the abstract and have been studied for their influence on the severity of ASD. *Lactobacillus rhamnosus GG* provides a good example. A review article relating to the effects of probiotics on cognitive function in 2019 was retrieved (8). The review highlighted the findings of a randomized controlled trial on the potential effect of *Lactobacillus rhamnosus GG* in reducing the risk of ASD development, and concluded that probiotics might be a safe and effective therapy targeting ASD. Since there are a variety of proposed beneficial health effects of probiotics, their use should be further explored for their potential benefits and side effects.

### Potential application of probiotics revealed by ProBioQuest: Epilepsy

A search of PubMed.gov articles under “probiotic” or “probiotics” and “epilepsy” turned up only 20 results from 2105 to 2021, indicating that the study of probiotics related to epilepsy is a new topic. One clinical study (“The Effect of Probiotic Supplementation in Drug-resistant Epilepsy Patients”) was recorded, and its related journal article (“The beneficial effect of probiotics as a supplementary treatment in drug-resistant epilepsy: a pilot study”) was published in 2018 (9).

This database not only archived probiotics-related journal articles, clinical trials, and patent records, but also records related to gut and gut microbiota. By searching “epilepsy “ and “gut”, we turned up 92 items that mention epilepsy and gut. From the column of publication year, only a few reports on the relationship between epilepsy and gut before 2015 were returned. However, after 2018, journal articles on that topic doubled in number. The studies of gut have increased as more researchers pursue the understanding of the gut and its ecological potential (10). From the Keywords Bank, the “Microbiome” category showed some genuses and species that have been studied or investigated, such as *Prevotella, Bacteroides, Clostridium difficile* and *Entamoeba histolytica*. These bacteria and their genomes can provide a starting point for study of possible probiotic strains with potential for reducing the frequency of seizures.

### Significance of ProBioQuest to health sectors

As consumers increasingly demand alternatives to pharmaceutical drugs (11), probiotics have attracted attention as possible biotherapeutic agents. Probiotics are commonly taken for the maintenance of health and wellbeing, alleviation of gastrointestinal discomforts and allergic conditions, and during antibiotic therapy to replenish the stock of friendly bacteria (12). It is evident that the probiotic market has been increasing in the past few years (13). However, patients who are interested in probiotics have complained that it is difficult to find reliable information on the functions and mechanisms of action of probiotics (14). Although health professionals are expected to handle enquiries about probiotic use from patients and consumers, they too have said that they need to educate themselves more about probiotics (11,15). Many health professionals let patients specify a particular probiotic strain to patients (16), perhaps because they are not confident in their own knowledge in this area. Since ProBioQuest allows health professionals to search for information specific to probiotic strains, it can make them more confident in prescribing or recommending probiotics. Books, expert magazines, and websites have been the key sources of information on probiotics acquired by health professionals (11), and keeping abreast of developments can be time-consuming. As ProBioQuest is an open, continuously updated system, users could explore up-to-date probiotic information with high relevance and credibility. Moreover, ProBioQuest can act as a learning program to raise awareness and knowledge on probiotics among current and future health professionals. By consulting the peer-reviewed academic papers in PubMed.gov, health professionals can obtain solid evidence-based knowledge that is appropriate to their clinical application. More importantly, the search results in ProBioQuest are classified, when acquired, in terms of probiotics, diseases, microbiome, food, original sources, and time period. ProBioQuest will help busy health professionals to save time in mining information and acquire a knowledge of probiotics quickly and efficiently.

ProBioQuest is a semantic tool for trends identification which helps members of the biomedical community to discover the unexplored clinical applications and knowledge gaps concerning the mechanisms of actions of probiotics. Apart from gastrointestinal system, probiotics are suggested to be applied in the field of weight management, respiratory, immune, skin, nervous, and cardiometabolic system (17). Although the health benefits of probiotics in research studies are increasingly being reported, no probiotic has yet been approved by the US Food and Drug Administration for prevention or treatment of health problems (18). This is understandable because of the inconsistent data between studies and inadequate standardized protocols (19) Nevertheless, research on probiotics or the microbiome could be sponsored by the National Centre for Complementary and Integrative Health (22), highlighting the importance of disease management through modulation of gut microbiota. Performing a literature review for funding applications using the traditional search engines can be time-consuming when screening a long list of miscellaneous reports. The use of ProBioQuest makes it quicker to pinpoint the necessary information. A broad spectrum of possibilities are offered, from simple keywords to sophisticated advanced searches. Besides limiting the set of relevant articles according to the key concepts, users could further improve the precision of information searching based on the types of database and articles, and their preferences. Additionally, they could export either selected or all search results, allowing them quickly and efficiently to save a collection of relevant information for an in-depth literature review.

A strong marketplace in probiotics is emerging along with the growing number of well-controlled human studies on probiotics. Probiotics have aroused widespread general interest owing to their potential to improve health and alleviate disease (20). Over 2000 clinical trials and 300 systematic reviews on probiotics were included in the biomedical database Medline in July 2019 (18). It is noteworthy that some established probiotics could be used together with pharmaceutical agents, foods, and lifestyles for health management (20). The value of the global probiotics market is estimated to increase from its 2020 value of USD 54,775.5 million to USD 95,252.9 million in 2028 (21)(Grand View Research, 2021). Since manufacturers aim to expand their market share of the probiotics market through new product innovations and frequent product launches, they have a responsibility to develop high-quality probiotic products supported by scientific evidence. Safe probiotic strains possessing relevant functional properties should be selected by manufacturers for human use. Therefore, ProBioQuest would serve as an important pool of up-to-date credible journal articles and clinical trials to assist manufacturers, especially those not familiar with scientific research in the field of probiotics, to evaluate and validate the beneficial effects of probiotics and formulate evidence-based probiotic products that can demonstrate clinical efficacy. By using our database, manufacturers can not only explore the technologies of promoting probiotic survival along the gastrointestinal tract and manufacturing process, but also the potential for their innovations to be patented.

## Limitations and further development

An examination of our data capturing and analyses processes has revealed certain limitations of ProBioQuest. Firstly, revealing strain numbers is difficult. Secondly, keywords may be missed. The format of bacterial strain numbers is not standardized in the literature. New strain numbers are generated constantly. We could not think of a way to capture strains into our keyword lists. Users would have to get the species first and the strain numbers would need to be identified in the paper. We tried to be as extensive as possible when we built the keyword lists for indexing the articles in ProBioQuest. However, new keywords appear quickly and many synonyms exist for many keywords, for example, as common names, which makes exhaustive keyword lists very difficult, if not impossible, to produce. We will try to address these limitations as we continue to develop ProBioQuest. We plan to set up a section in the web interface to invite suggestions on keywords and for users to report new probiotics and microbiome bacterial strain numbers, so that we can add them to the keyword lists.

## Conclusion

Probiotics are used to supplement medical treatments in many diseases and health conditions. Interest in probiotics has increased tremendously in the past decade, and new articles, records of clinical trials and details of patents granted are published continuously. Despite this growing torrent of information, dedicated collections and analytical services have so far been lacking. The few available databases are either very small in scale or not accessible. We have constructed ProBioQuest, a database with probiotics as the central theme, with semantic analytical functions, external links, and a user-friendly web-based interface. ProBioQuest is easy to use and captures up-to-date articles from PubMed, Clinical trials.org, and PatentsView.org automatically. This tool meets the needs of probiotics information gathering and analysis for the probiotic scientific community, industry, and health professionals. More than 2.8 million articles were collected by the end of 2021. Automated information technology-assisted procedures enable us to collect data continuously, providing the most up-to-date information available. Statistical functions and semantic analyses are provided on the website as an advanced search engine, which makes this database a valuable semantic tool for information searches and analyses. Users can identify trends, discover hubs, identify popular, emerging, and potential applications, and make in-depth content analyses.

## Data Availability

All data produced in the present study are available upon reasonable request to the authors

## Acknowledgements

We would like to thank Probiolife limited, Ms. Shally Wong and sifa Wang for their support, and Dr. David Wilmshurst for editing the English in this manuscript.

